# Impact of cannabis mass gathering events on mental health and health service utilization

**DOI:** 10.1101/2020.04.20.20073387

**Authors:** Patrick Lombardo, Andrew Lim, Andrea A. Jones, Daniel Vigo, William G. Honer, Jennifer Duff, G. William MacEwan, Fidel Vila-Rodriguez

**Author notes:** Correspondence to: Assistant Professor, Michael Smith Foundation for Health Research Scholar, Department of Psychiatry, University of British Columbia, Director, Non-Invasive Neurostimulation Therapies Laboratory & Schizophrenia Program NINET.CA / Twitter: @NINETLAB, Detwiller Pavilion, 2255 Wesbrook Mall, Vancouver, British Columbia, V6T 2A1. <u>Competing interests:</u> Dr. Lombardo reports grants from the Swiss National Science Foundation (P2LAP3_164907). Dr. Vila-Rodriguez reports research grants from CIHR, Brain Canada, Michael Smith Foundation for Health Research, and Vancouver Coastal Health Research Institute; reports receiving in-kind equipment support for this investigator-initiated trial from MagVenture; and has been on an advisory board for Janssen. The remainder authors declare that they have no competing interests. <u>Funding:</u> There was no funding source for this study.

## Abstract

**Background:** The Canadian government has legalized and regulated access to cannabis as of October 2018. In this context, there is a need to analyze data that may provide insights on the effects of increased accessibility and tolerance for cannabis use. One source of data is the phenomenon known as “4/20”, a decades-old yearly mass gathering event supporting the legalization of cannabis. These events offer naturalistic epidemiologic data to ascertain specific impacts of cannabis consumption in a context of increased tolerance on health service utilization. Our study assessed the association between cannabis mass gathering events and health service utilization related to mental illness and substance use disorders at the nearest local emergency department.

**Methods:** Emergency department service utilization data (2005-2015) was used. The sample analyzed consists of emergency department visits due to mental and substance use disorders. A multiple linear regression model was used to predict the number of daily visits with year, month, day of the week, and day of income assistance distribution as independent variables. Daily residuals were averaged, and residuals for the days with the highest number of visits were compared with the mean residual number of visits. Also, correlation of number of visits with attendance to mass gathering events was explored.

**Results:** The residual number of visits for mental health and substance use disorder was the highest on April 20^th^ 2015 (n=51.0, z-score=11.0, p<0.001), and on days associated with subsequent cannabis mass gathering events. Moreover, this number of visits is positively correlated with the number of attendees at the “4/20” event (Pearson’s correlation coefficient: 0.76, 95% CI: 0.19 to 0.956, p=0.002), and increased over time.

**Conclusion:** Cannabis mass gathering events were associated with an increased number of emergency visits for patients with mental health and substance use diagnoses at the nearest local emergency department. In the context of legalization and regulation of cannabis use, these specific gatherings will not necessarily be discontinued. Indeed, as per news reports the recent post-legalization “4/20” drew tens of thousands of people in Vancouver. Also, in the new context other non-specific mass gatherings may also lead to foreseeable episodic surges in ER utilization. In light of this and from a public health perspective, services need to be prepared to care for predictably larger numbers of people suffering cannabis intoxication during mass gatherings, as well as to make provisions to provide all other services that are regularly needed for other emergency conditions. Also, educational campaigns about responsible use during these events will become particularly important, as well as offering on-site support, triage and basic services. This will allow for specific care to be provided in a non-stigmatizing manner, proportional to need, and without overcrowding general emergency services.

## 1. Introduction

Cannabis is one of the most commonly consumed non-prescribed drugs in the world (1). Several stake-holders and academics suggest that regulated legalization of cannabis is less harmful than strict prohibition, but the health and health services implications of this policy are not thoroughly known (2–9). The consumption of cannabis is associated with different health effects, including acute responses such as impaired short-term memory, decreased motor coordination, and acute psychosis. Chronic effects may include increased risk of addiction, alterations in brain development, and psychosis (10–12). Notably, adolescents may be particularly sensitive to adverse health effects (13,14).

In Canada, consumption of cannabis by adults has been legalized in October 17, 2018. British Columbia appears to be the Canadian province with the highest availability, consumption, and acceptance of cannabis (15). For decades, mass gathering events at which cannabis consumption is accepted have been organized by people supporting more widespread access to cannabis. The most popular of these events, “4/20”, has occurred on April 20^th^ each year since 1995, (16–18). Of note, starting in 2011 the Vancouver event developed a commercial component, whereby vendors offer cannabis merchandise associated with consumption (19,20). The attendance has consistently increased, this year estimated by news organization in the tens of thousands; of note, in 2015-the last year of data analyzed here-it gathered 25,000 participants, a greater than 20-fold increase from 1997 (20,21). Cannabis mass gathering events have emerged elsewhere in Canada, USA, and cities around the world (22–26). These mass gathering events offer a naturalistic opportunity to ascertain the impact of acute cannabis consumption on both service utilization and health outcomes. In this regard, recent work with data from the US reported the “4/20” event was associated with increased rates of fatal motor vehicle accidents (27).

We sought to determine the relationship between cannabis-related mass gathering events and temporally- and geographically-proximal emergency health service utilization over ten years using anonymized administrative data. We hypothesized that cannabis-related mass gathering events were associated with significant increases in local emergency health service utilization. Specifically, we investigated the correlation between the number of people attending the event and the number of health service utilization visits associated with mental health and substance use disorders at the nearest local emergency department.

## 2. Methods

The study was approved by the clinical research ethics board at the University of British Columbia and Providence Health Care.

### 2.1. Data source

Anonymized data provided by Providence Health Care including the visits to the emergency department of the St. Paul’s Hospital in Vancouver from April 2005 to December 2015 were used (28). This hospital hosts the closest emergency department to the area where the cannabis mass gathering events have taken place (Appendix A). For each emergency department visit, the database included the sex and age of the patient, date of consultation, chief complaint, and physician discharge diagnosis.

### 2.2. Study sample and variables

The full dataset included all emergency department visit between 2005 and 2015. Other than for the descriptive demographic section, we focused our analyses on a sub-sample that consisted exclusively of emergency visits related to mental and substance use disorders. 982 such visits were identified through physician discharge diagnoses including codes F10 to F19 of ICD-9 and ICD-10, the standard method used in British Columbia’s hospital system. The F12 code (cannabis-related disorders) was used to identify visits associated with cannabis consumption. The “physician discharge diagnosis” results from the discharge process: at the time of discharge, a physician codes the patient’s discharge diagnosis using the International Statistical Classification of Diseases and Related Health Problems (ICD).

The 9^th^ revision (ICD-9) was used before July 27^th^ 2005, and the 10^th^ (ICD-10) revision thereafter. Overall,

The “chief complaint” (used in the sensitivity analysis) is determined upon arrival to the emergency department by the triage nurse. Each visit was recorded and classified into a category belonging to one of 18 different existing systems, including “mental health” and “substance misuse”.

Predictors used in the multiple linear regression model were year, month, day of the week, and day relative to income assistance distribution, to capture the time related variables that potentially determine daily ER visit variation.

In order to estimate the number of participants at the “4/20” cannabis mass gathering event, and in the absence of official data sources, contemporary news publications and a cannabis counterculture website were explored. The source of these estimates was not systematically declared, and these were likely based on the observations of the organizers or journalists (Supplemental Table 1) (16,18,19,21,29–33).

### 2.3. Statistical analysis

First, the daily number of visits with an ICD diagnosis related to a mental health or a substance use disorders was extracted for the entire time period. Second, multiple linear regression analysis was used to estimate the number of daily visits by year, month, day of the week, and day relative to income assistance distribution (usually the last or second last Wednesday of each month). The residual daily number of visits can be interpreted as the variance in daily visits not explained by these temporal factors. Third, the residual daily visits for several mass gathering events (i.e., 4/20, Canada Day, Halloween, and New Year’s Eve) were compared to the mean residual number of visits. The null hypothesis was that the residual observed number of visits on a given day is not different from the mean residual number of visits. The strength of the relationship between the residual number of visits, the number of attendees at the 4/20 event, and the year were measured using Pearson’s correlations ®. This approach was used to explore a subgroup of patients with an F12.0 code (cannabis-related disorders) visits. The results of the main analysis were confirmed with a sensitivity analysis using the chief complaint instead of the ICD discharge diagnosis.

The analyses were performed using R version 3.3.3 (34) with the following packages *car* (35), *ggplot2* (36), *ggrepel* (37), *plyr* (38), and *zoo* (39).

## 3. Results

### 3.1 Demographics

From April 1^st^ 2005 to December 31^st^ 2015, the cumulative number of visits at the emergency department was 747,321. The number of visits increased year by year: there were 57,916 visits in 2006, and 83,630 in 2015. The daily mean (standard deviation, SD) number of visits for mental health and substance use disorders was 18.0 (6.2) and represented 9.4% of total visits. An upward trend in the number of visits for mental health and substance use disorders was observed over the ten-year period. The ICD discharge diagnosis was missing in 19.8% of visits. Patients with visits with complete data were similar in age (mean 44.8, SD 18.4, in the complete data and 44.6, SD 18.5, overall), gender (40.6% and 40.5% of female, respectively), and in the rate of visits with a chief complaint related to mental health and substance use disorders (10.1% and 9.8% respectively).

### 3.2. Residual number of visits

Across the ten-year period, a greater number of visits with an ICD diagnosis related to a mental health or a substance use disorders was measured from April to October, from Thursday to Saturday, and on the four days after income and disability assistance was issued. The residual number of visits was obtained after adjustment for these parameters and refers to the residual number of visits for mental health and substance use disorders. The day with the highest residual number of visits was Monday, April 20^th^ 2015. On that day, there were 51.0 more visits than expected (z-score: 11.0, p<0.001). The top four days with the highest observed or residual number of visits were associated with a cannabis mass gathering event (Table 1). The five dates associated with the highest mean residual number of visits were: April 20^th^, January 1^st^, July 1^st^, October 31^st^, and November 1^st^. Figure 1 provides an overview of the residual number of visits on these dates. From 2010, the residual number of visits on April 20^th^ of each year was constantly higher than the mean, and this number was significant from 2013 and onwards (2013 n=31.8, z-score: 6.9, p<0.001) (Table 2).

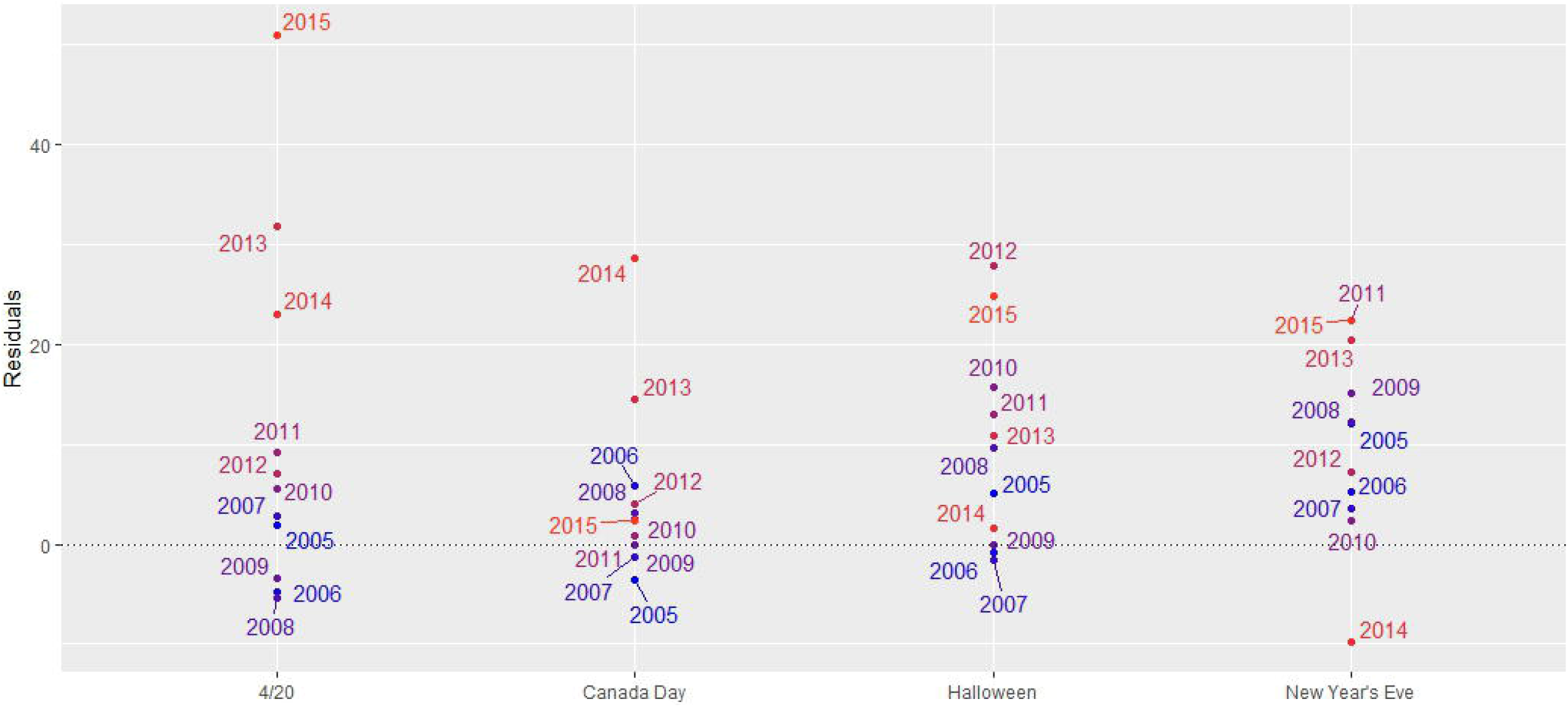

### 3.3. Correlation Analyses

Year after year, there was an increase in the number of attendants to the “4/20” event (r: 0.94, 95% CI: 0.72 to 0.99, p<0.001). Furthermore, there was a statistically significant positive correlation between the number of attendees of the event and the residual number of visits (r: 0.76, 95% CI: 0.19 to 0.956, p= 0.02). In contrast to the dates of January 1^st^, July 1^st^, October 31^st^, and November 1^st^, the residual number of visits increased significantly from year to year on April 20^th^ (r: 0.82, 95 CI: 0.44 to 0.95, p=0.002) (Supplemental table 2).

### 3.4. Cannabis-Related Disorders

The number of visits with cannabis-related disorders (F12 code) on April 20^th^ from 2011 to 2015 roughly matched the residual number of visits on the same dates (Table 2). July 1^st^ 2014 was associated with a cannabis mass gathering event: there was 28.7 residual number of visits and 20 visits with cannabis-related disorder. The number of visits in the top five of the days with the highest residual number of visits for cannabis-related disorders was significant and was associated with cannabis mass gathering events (Supplemental table 3 and supplemental figure 2). The F12.0 (cannabis-related disorders) code from the ICD-10 was used in about one-quarter of the cases on the April 20^th^. Finally, the cannabis-related disorders code (F.12.0) was assigned to 29 visits involving minors (< 19 years old) on April 20^th^ (Table 3). Most of the patients receiving an F12.0 diagnosis on April 20^th^ had a chief complaint registered at triage that was compatible with acute intoxication with cannabis (Supplemental table 4).

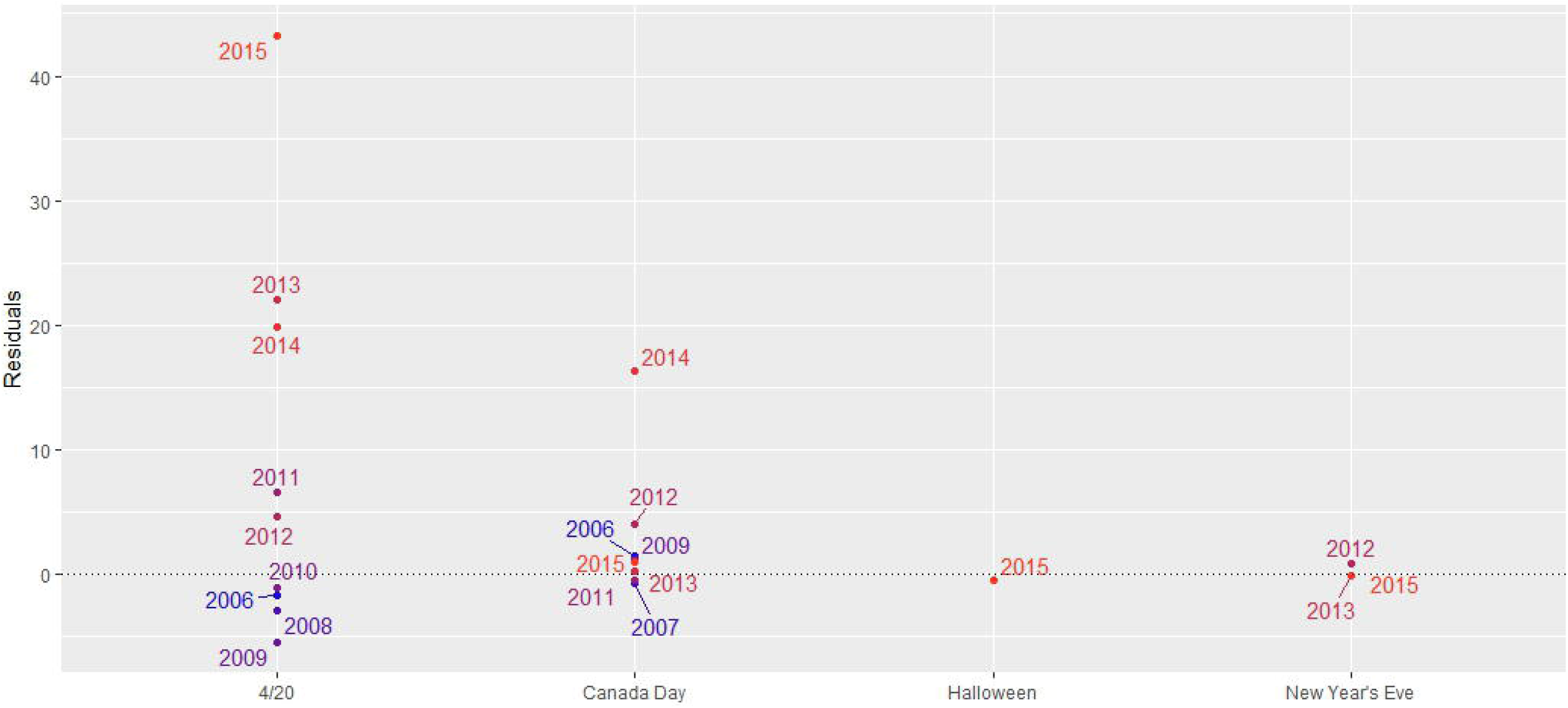

### 3.5. Sensitivity analysis

A chief complaint was registered for 96.0% of the visits for any reason, and 9.8% of these visits had a chief complaint classified in the mental health or substance use disorders system. In 70.2% of these visits, a final ICD diagnosis from the mental health or substance use disorders was retained. The results obtained from this analysis were comparable to those obtained with the main analysis (Supplemental table 5 and supplemental figure 3).

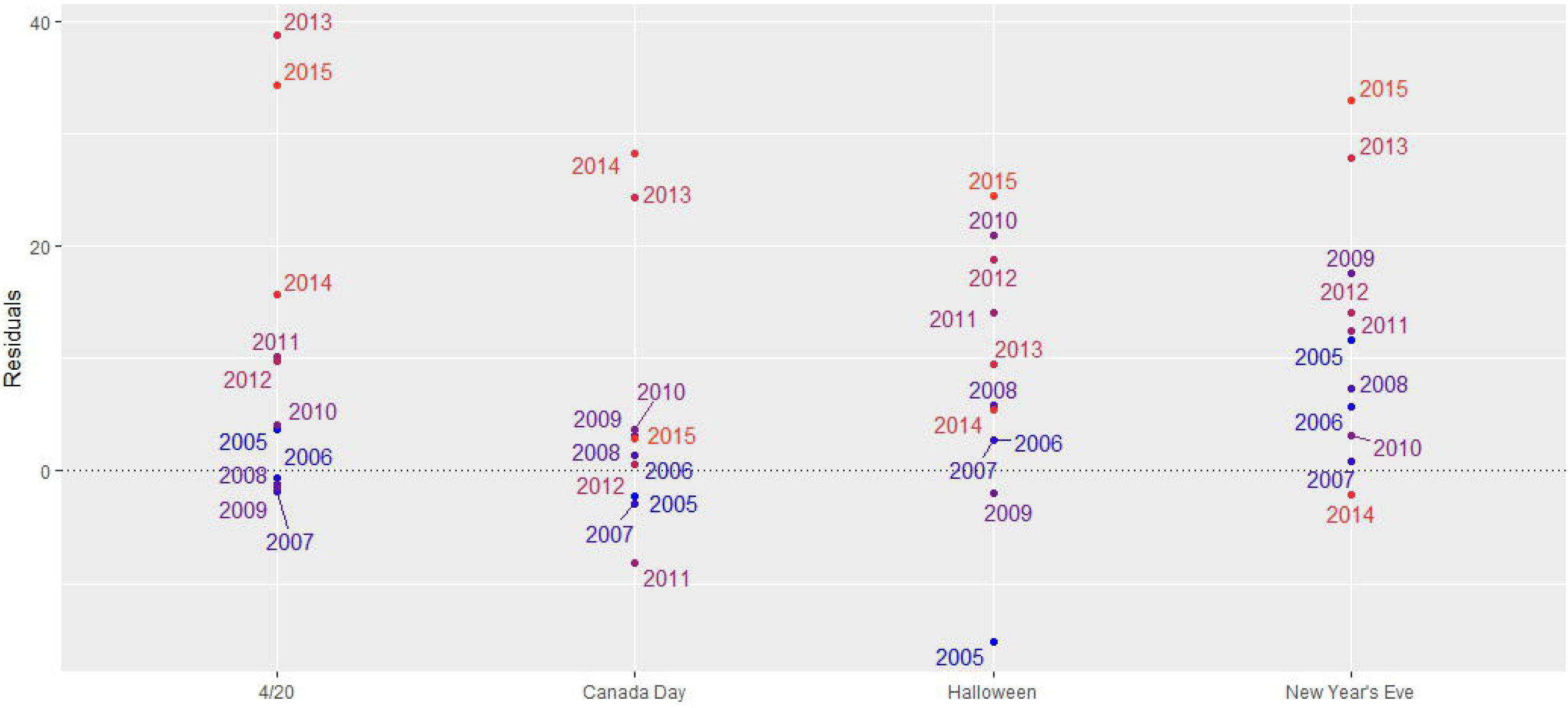

## 4. Discussion

This is the first study to ascertain the impact of cannabis mass gathering events on the frequency of health care emergency services utilization. The findings highlight the impact of the “4/20” event that takes place each year in Vancouver on April 20^th^, on local health service utilization due to mental health and substance use disorders. Specifically, from 2010 to 2015 there was a higher residual number of visits for mental health and substance use disorders at the emergency department of St. Paul’s Hospital on that date. This increased number was statistically significantly different from the mean residual from 2013 onwards, coinciding with a significant increase in attendance at the event.

April 20th was among the days with the highest residual number of visits (three of the top four dates). July 1^st^ 2014 saw the third greatest number of visits, coincident with another cannabis mass gathering event (Canada’s national holiday was chosen for another event in support of cannabis legalization, named “Cannabis Day”(40)). The residual number of visits was not increased on July 1^st^ 2015, which coincided with a cannabis mass gathering event that was not authorized by the authorities (41,42). The increase in visits on April 20th correlated strongly with the increase in attendance at the “4/20” event, pointing to a very plausible effect of number of attendees and number of people using cannabis on number of cannabis related medical emergencies. However, it should be noted that the period 2005 to 2010 resulted in an increase in attendance, but this did not impact strongly on the number of residual visits. Notably, in 2011 the event increased the presence of booths selling cannabis-related products, and this may have promoted cannabis consumption. The specific ICD-10 diagnosis for cannabis-related disorders (F12) was used in about one-quarter of the cases on April 20^th^, and from 2011, the number of visits on that day with this diagnosis approximately corresponded to the residual number of visits. These results provide a strong association between the greater number of visits on April 20^th^ and the 4/20 cannabis event. The greater number of minors receiving an F12 diagnosis on April 20^th^ also supports this conclusion.

Our study had several strengths: two different systems of classification to categorize the precipitants of the visits were used and analyzed (discharge diagnosis and chief complaint at intake), and data from a 10-year period were available for analysis. However, an ICD diagnosis was missing in 19.8% of the total visits, which could lead to bias. This limitation could be mitigated by the sensitivity analysis that used the chief complaint system entry, that was available for 96.0% of the visits, and confirmed the results of the main analysis. Another limitation is that no publicly available official documents register attendance during the event. Also, since data from hospitals outside of St. Paul’s were not available, we cannot infer if there was a net increase in the number of visits in Vancouver and surroundings, or whether there was simply a redistribution of visits from other hospitals to hospitals near the 4/20 event.

Finally, an overutilization of the F12 code on April 20^th^ and an underutilization the rest of the year is very likely: as the consumption of cannabis was tolerated during Cannabis Day and prohibited during the other days, it could be possible that consumption of cannabis was more likely to be reported by emergency department visitors on the cannabis related events and concealed on the other days. Similarly, health providers may also be biased to attribute symptoms to acute cannabis use on April 20^th^. To lessen this bias, different measures were taken: first, the main analyses were performed on the visits for mental health and substance use disorders and not on the specific F12 code. Second, the concordance between the F12 diagnosis used on April 20^th^ and the chief complaint description registered at the triage was compared.

These findings are far from trivial. Our data support the notion that concentrated mass consumption of cannabis in a tolerated context is associated with increased emergency service utilization. This has two potentially negative effects: the health consequences of intoxication themselves and the saturation of emergency rooms, potentially affecting service to other patients. Our findings indicate that emergency service needs due to cannabis consumption in the context of mass gatherings predictably increase, providing an opportunity for the health system to prevent some of the negative consequences. For example, dissemination of information on responsible utilization, control mechanisms on utilization by minors, and education on signs and symptoms of intoxication as well as on vulnerable population (minors, pregnant women, people with or at increased risk of psychotic disorders) may prevent or mitigate a fraction of the damage. These vulnerable groups may require increased regulation, prevention or care, as research indicates that the people at increased risk for psychosis and people undergoing neurodevelopment may be particularly sensitive to the effects of cannabis consumption (13,14). Further, on-site provision of triage, support and referral for emergency department visit only when medically needed may have two desirable consequences. First, on-site services may be delivered in a non-medical setting by well-trained community providers and peers. This would have positive cost implications as well as potentially derease stigma and increase acceptability of information and services. Second, it would free up valuable and costly emergency department resources, which would still be available when required.

In summary, these findings offer insights into the increased demand for mental health and substance use related emergency services during mass gatherings in a context of increased utilization, and indicate potential approaches to improve services delivered to people who use cannabis and other users in general.

## Data Availability

Due to privacy laws data is not available

## Acknowledgement

This paper is dedicated to the memory of Dr. Elliot Goldner.

